# A Large Language Model-based Approach for Analyzing Covariates of Health Equity in Registered Research Projects

**DOI:** 10.1101/2024.09.24.24314327

**Authors:** Navapat Nananukul, Mayank Kejriwal

## Abstract

Large language models (LLMs) have made significant advancements in natural language processing, offering broad applications in multiple domains. This study explores the use of the GPT-3.5 LLM to conduct efficient and robust computational analysis of registered research projects on the *All of Us* platform. Specifically, we explore the association between projects pursuing health equity research and: the project’s use of demographic categories (which *All of Us* enables), the multi-institutional composition of the team leading the project, and the involvement of R2 institutions (compared to only R1 institutions). We demonstrate the utility of GPT-3.5 in automating tasks ranging from generating Python scripts for extracting attributes from free text (such as project description and goals) to identifying and classifying institutions as R1 and R2, and summarizing project details into Unified Medical Language System (UMLS)-coded medical keywords. These contributions significantly reduced manual workload, allowing researchers to focus on more in-depth analysis. Our results reveal health equity insights not readily available in the original *All of Us* research hub. Specifically, we find a strong positive association between the use of demographic data and projects focused on health equity, while other associations such as health equity projects conducted by institutions were positive but weaker and more dependent on specific project topics.

## Introduction

Health equity has been recognized as a significant concern in the United States [1-5], with Whitehead framing it in the early 1990s as “equal access to available care for equal need, equal utilization for equal need, equal quality of care for all” [6]. A framework created in the 1990s by the National Institute on Minority Health and Health Disparities similarly provides comprehensive definitions of heath equity and emphasizes the importance of addressing factors to reduce health disparities [7].

The *All of Us* research platform is an ambitious initiative that envisions improving health equity (as one of its goals) by “catalyzing” an ecosystem of stakeholders in gathering and analyzing data from more than one million individuals residing in the United States [8]. Since its launching, the platform has witnessed many projects being registered and covering topics ranging from mental health to cardiovascular disease [9]. Registered projects are briefly annotated with both structured data (including *demographic variables*, if any, being used in the project) and more unstructured text data, such as *anticipated findings*.

Properly studied, these projects can be used to answer a rich set of open sociological questions involving covariates of projects that explicitly choose to address health equity as one of their goals [10-12]. In this article, we systematically analyze hundreds of registered *All of Us* project descriptions and associated metadata, across five broad topical clusters, to investigate the odds of a project addressing health equity as a goal and the project, (i) being led by a multi-institutional team (suggesting interdisciplinary research), (ii) making use of demographic variables (suggesting the value of *All of Us* in providing fine-grained data that facilitates health equity studies), and (iii) involving doctoral Carnegie-classified R2 universities (“high research activity”) compared with R1 institutions (“very high research activity”) [13], which would suggest greater institutional diversity in exploring health equity.

Our study contributes to a growing body of research on the prevalence and drivers of health equity projects currently being pursued in the United States and several other countries [14-17]. Some of this research is policy-driven, while others focus on the importance of health equity in implementation science. However, a systematic and quantitative review of registered projects that places health equity as a first-class subject of analysis has thus far been lacking. Because they involve personnel and resources, and are formulated with good-faith *intent*, such projects should be studied independent of whether (or not) they lead to tangible *impacts* like publications. Historically, data for such endeavors was not available but has since become so, because of platforms like *All of Us*. We choose to analyze these projects directly, including bona-fide descriptions of anticipated findings and *a priori* scientific questions of interest when registering the study.

We note that conducting such a study at scale is methodologically non-trivial because *All of Us* does not directly code for variables like health equity, variables flagging multi-institutional teaming, and whether an institution is an R1 or R2 university. All of these are required for testing the hypothetical associations described earlier and must be inferred for hundreds of projects using efficient and largely automated processes. In *Methods*, we provide a detailed pipeline that makes use of both a comprehensive medical vocabulary like the Unified Medical Language System (UMLS) and a large language model (LLM) like GPT-3.5 for text analysis, coupled with judicious amounts of manual annotation and verification, for obtaining key variables of interest.

### The *All of Us* Research Hub

The *All of Us Research Hub* is a platform that hosts a large collection of health research projects from a diverse participant population across the U.S. It serves as a vital resource for researchers, providing access to over 13,000 registered medical projects. Each project contains detailed information, including research questions, purposes, approaches, anticipated findings, dataset types, and team details. The platform’s interactive data browser allows users to perform keyword searches, explore project data snapshots, and view comprehensive research descriptions, making it a potentially valuable tool for large-scale health equity research.

In this study, we utilized the *All of Us Research Hub* as our primary data source to investigate medical research projects. Figure 1 illustrates the platform’s interface, displaying a list of projects from the search results for the keyword “Diabetes”. We also show the detailed project information that is available when users select a project. While the hub offers a robust tool for exploring individual projects, some information needed for the analysis may not be available to researchers. For example, in considering the organizations that members of a project’s research team are affiliated with, All of Us does not code for the Carnegie classification of an academic organization (e.g., R1 versus R2). Moreover, conducting large-scale manual analysis of thousands of projects requires additional preprocessing steps which can be time-consuming and distracting from the main analysis. Therefore, we used GPT-3.5, a large language model, to assist with data extraction tasks such as identifying institutional affiliations and extracting health equity-related keywords from the unstructured project descriptions. By automating these processes, we efficiently processed and analyzed medical projects across five targeted medical keywords, enabling a comprehensive analysis of the projects registered in the platform.

**Figure 1.**
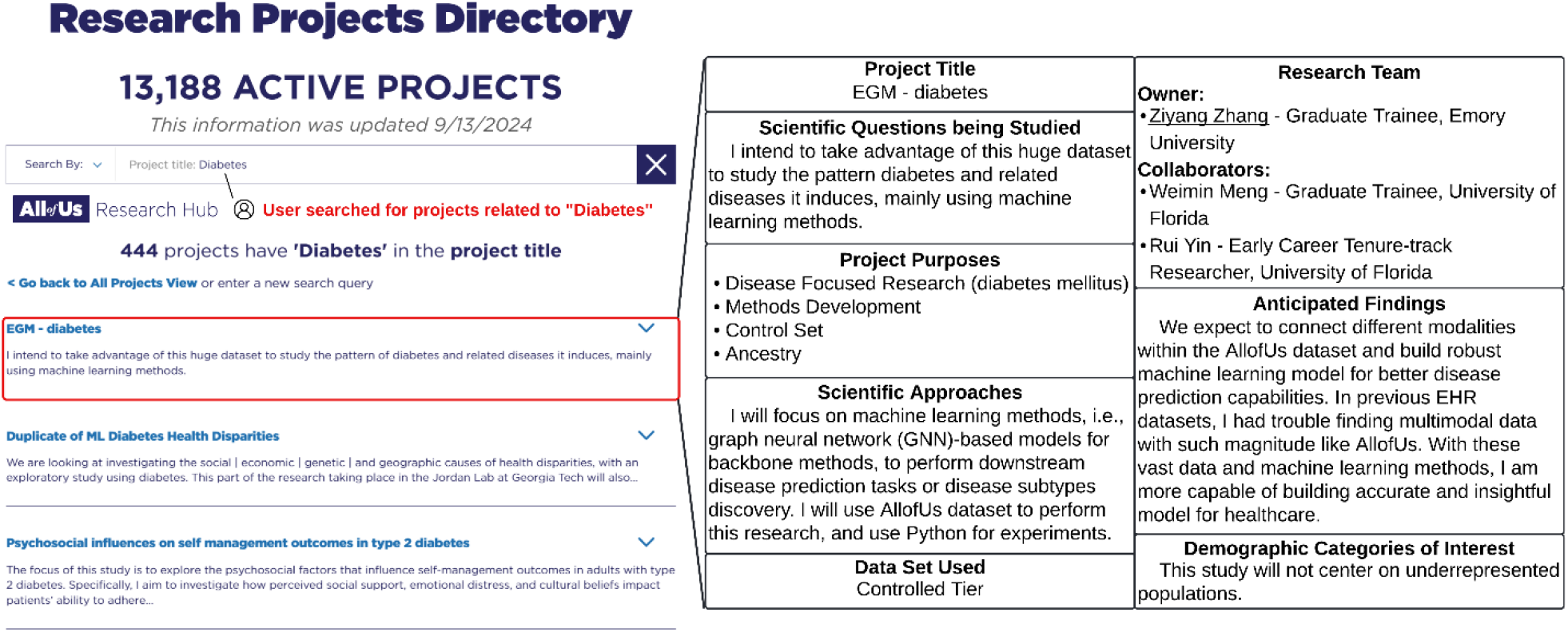
An illustration of the *All of Us Research Hub* interface. The left image displays search results for projects related to the keyword “Diabetes,” highlighting how researchers can utilize the search bar to find specific projects. The right image displays detailed information about the project that users selected.

## Materials and Methods

Because *All of Us* does not directly code for variables like health equity, variables flagging multi-institutional teaming, and variables indicating whether an institution is an R1 or R2 university, such variables must be inferred for hundreds of projects using the unstructured text descriptions and (where applicable) external data sources. We constructed such a pipeline (Figure 2) that makes judicious use of a large language model (LLM) like GPT-3.5 [18] for text analysis and coding of variables that traditionally required painstaking manual effort [19-20] and that may implicate potential cognitive biases [21-23].

**Figure 2.**
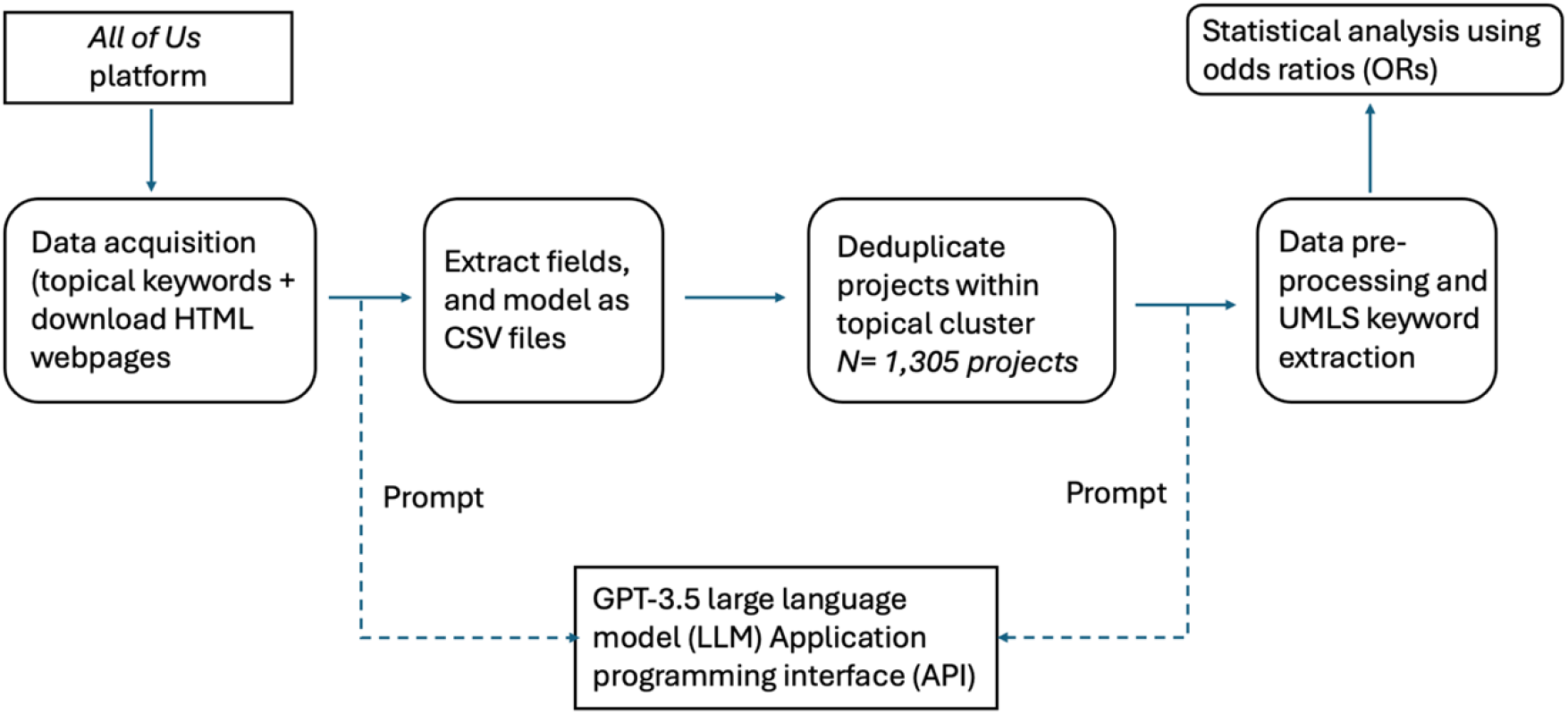
A simplified workflow illustrating data acquisition (from the *All of Us* platform), preprocessing, deduplication, and data augmentation (e.g., UMLS keyword extraction) steps. Code and GPT-3.5 prompts underlying these modules are discussed in the main text.

### Data Acquisition and Field Extraction

We acquired data for the study from the *All of Us* platform on five topics of broad interest: *asthma / pollution, cardiovascular disease, diabetes, dementia / Alzheimer’s*, and *mental health*. For each topic, we used simple keywords against the *scientific questions being studied* field to compile a topic-relevant list of projects, and retrieved a set of HyperText Markup Language (HTML) files for each subject from the search results, containing project details such as *titles, goals and aims, approaches*, and *team members*. Next, we extracted data from the HTML into Comma Separated Values (CSV) spreadsheets using a data extraction program based on a widely used web-text extraction Python library [24]. We used GPT-3.5 to assist us in writing the script, but manually verified the quality of the script. We also conducted checks to detect missing values, nulls, or blanks. Key descriptive statistics are provided in Table 1.

**Table 1.** Descriptive statistics on *All of Us* research project descriptions used in this study. The methodology for deduplication, and inferring additional fields from the raw text, such as the institutions and individuals, is detailed in the main text.

| Topic | Search keyword(s) | # registered projects (deduplicated) | # of unique individuals listed as team-members | Average number of individuals per team | % projects using demographic categories for study | # of unique institutions |
| --- | --- | --- | --- | --- | --- | --- |
| Mental Health | "mental health" | 244 | 365 | 1.82 | 63.52% | 140 |
| Dementia and Alzheimer's | "dementias", "alzheimers", "dementia" | 174 | 246 | 1.80 | 43.68% | 88 |
| Cardiovascular disease | "cardiovascular" | 388 | 588 | 1.86 | 50.00% | 153 |
| Asthma and Pollution | "asthma", "pollution" | 92 | 128 | 1.67 | 45.65% | 60 |
| Diabetes | "diabetes" | 407 | 600 | 1.91 | 51.84% | 176 |

### Deduplication of Projects

Next, we identified duplicate entries within our datasets from the compiled CSVs. Duplicates are due to two primary causes: (1) instances of projects being registered multiple times on *All of Us* (e.g., by different members of the same team), and (2) the occurrence of the same project under multiple search keys due to use of related search keys e.g., many projects will overlap when independently using “dementia” and “alzheimers” as search keywords for retrieving results. We used a thresholded *bag-of-words* approach from the text analysis community frequently used in the deduplication of semi-structured data [25], a simple implementation of which is provided (see *Code Availability*). Manual inspection of randomly sampled project pairs after running and tuning the algorithm showed that deduplication could be achieved with 100% accuracy due to high text- and field-overlap in such project pairs.

### Preprocessing and Keyword Extraction

We performed data preprocessing, both to extract more fine-grained information from project descriptions, and to infer additional fields from the original dataset. The former involved determining whether the project was being led by members of more than one institution (multi-institutional team), whether (and what) demographic variables were used, and total numbers of unique institutions and individuals. To determine multi-institutionality, we first concatenated the team members, with their roles and institutions, as a text-string. Because this field does not have a fixed structure and cannot be parsed using a simple heuristic function, we elected to prompt GPT-3.5 on each concatenated string to extract three lists of (i) team member names; (ii) corresponding roles; and (iii) affiliated institutions. Using (iii), we further prompted GPT-3.5 to classify each institution on whether it is R1, R2, or neither. We manually sampled a small set of projects and metadata to verify the complete accuracy of this step. Figure 3 (top and middle) illustrates this process, showing how GPT-3.5 extracts structured information from unstructured project descriptions and classifies institutions into R1, R2, or neither.

**Figure 3.**
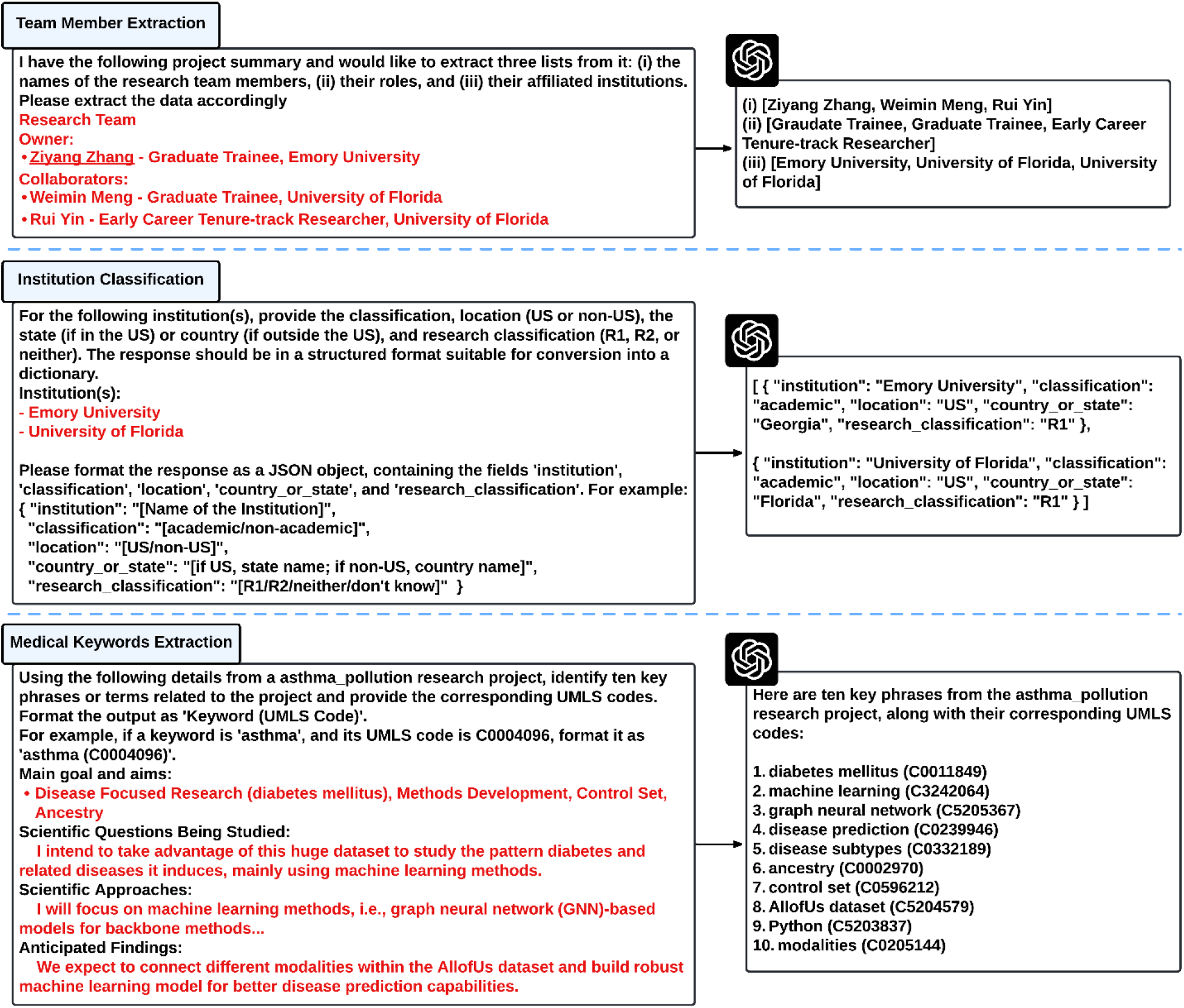
Three examples demonstrate how GPT-3.5 assists in data extraction and generation from unstructured text. In the first example (top), GPT-3.5 extracts the research team members, their roles, and affiliated institutions from an unstructured research team. In the second example (middle), GPT-3.5 processes a list of institutions (in red text) and returns each institution’s classification (academic or non-academic), location, and Carnegie classification (R1, R2, or neither). In the third example (bottom), GPT-3.5 extracts relevant medical keywords and their corresponding UMLS codes from the *All Of Us* project descriptions, as provided in the prompt instructions.

The key inferential field we sought to derive from the original data was a flag for whether the project had health equity as at least one of its goals. To do so in a robust way, we first provided GPT-3.5 with four fields (per project): *aims and goals, questions, approaches*, and *findings*. Next, we prompted it to extract the top 10 medical keywords using UMLS codes [26]. This yielded the top UMLS keywords each project is best related to. As shown in Figure 3 (bottom section), GPT-3.5 efficiently extracted UMLS-coded keywords, which were then used to flag projects for health equity relevance. Subsequently, we prompted GPT-3.5 by generating a comprehensive list of keywords related to “health equity,” recognizing that projects might address this concept in various ways. In total, GPT-3.5 provided a list of 42 keywords, such as “social determinants of health,” “health disparity,” and so on (see *Data Availability*). Any project flagged with at least one of these keywords was designated as pursuing health equity as one of its goals.

### Ethics Statement

The study only conducts analysis of registered publicly available scientific projects on the *All of Us* platform and does not meet the definition of human subjects research.

### Analysis

To investigate associations, we primarily use odds ratio (OR) analyses. All ORs are computed for individual topics, but where applicable, we report pooled ORs using fixed-effects Mantel-Haenszel pooling of ORs computed across all five topics. Based on the objectives stated earlier, we independently consider the association between a project pursuing health equity as a goal (outcome) and: (i) use of demographic fields, (ii) involving a multi-institution team, (iii) involving at least one R2 institution (with R1-only teams or institutions as baseline). For computing the OR in (iii), we exclude projects not affiliated with either R1 or R2 institutions, which results in a pruned dataset of 1,131 projects out of the original 1,305. To analyze co-occurrence strengths between keywords, we create a co-occurrence matrix that records the frequency of keyword pairs appearing together within projects. Using this matrix, we selected top 10 keyword pairs with the highest co-occurrence frequency across each of the five medical topics, illustrating the connection of keywords within and across topics. We visualize the co-occurrence using a chord diagram [27].

## Results

Our key statistical findings are provided in Table 2. We found significant association between use of demographic variables and health equity across all project topics (Mantel-Haenszel OR=4.069, 95% CI 2.719 to 6.087), ranging from 73% higher odds in the *dementia* topical cluster (OR=1.73, 95% CI 0.79 to 3.78) to 6.85x higher odds (OR=6.85, 95% CI 4.08 to 11.50) in the *cardiovascular* cluster. Using Cochran’s Q test, the null hypothesis for homogeneity of odds was rejected (Cochran’s Q = 11918.75, P=0.00), suggesting that the topic potentially modifies the strength of the effect, and that the pooled estimate above should be treated with caution. The magnitudes of OR effects are substantive, and except for *dementia*, they were all significant at the 95% confidence level.

**Table 2.** Associations, using odds ratios (ORs) with 95% confidence intervals (CI) between projects (within a topic) pursuing health equity as a stated goal and the project, (i) being led by a multi-institutional team versus single-institutional team (Column 2), (ii) making use of demographic variables versus not making use (Column 3), and (iii) involving at least one doctoral Carnegie-classified R2 university versus R1 institutions only (Column 4). Additionally, we use *, ** and *** to denote significant difference of the OR from unity at the 90, 95, and 99 percent confidence levels, respectively.

| Topic | Multi-institutional team | Demographic variable use | At least one R2 university |
| --- | --- | --- | --- |
| Mental Health | 2.173* (0.935, 5.050) | 3.635*** (2.086, 6.337) | 1.296 (0.674, 2.493) |
| Dementia and Alzheimer's | 1.481 (0.449, 4.894) | 1.729 (0.791, 3.778) | 0.564 (0.181, 1.776) |
| Cardiovascular disease | 0.883 (0.448, 1.743) | 6.846*** (4.076, 11.498) | 1.746* (0.983, 3.102) |
| Asthma and Pollution | 0 | 5.500*** (1.926, 15.706) | 1.735 (0.292, 10.301) |
| Diabetes | 1.495 (0.862, 2.591) | 3.606*** (2.309, 5.631) | 1.886** (1.154, 3.092) |
| Mantel-Haenszel pooled OR | 1.377 (0.821, 2.307) | 4.069 (2.719, 6.087) | 1.522 (0.979, 2.366) |

There is only slight evidence of higher odds of multi-institutional team projects addressing health equity as a goal, except for *cardiovascular* and *asthma*. The test for homogeneity was again rejected, suggesting the modifying effect of the topic on each association. For *mental health*, the OR was 2.17 (95% CI 0.94 to 5.05), for *dementia* the OR was 1.48 (95% CI 0.45 to 4.89), and for *diabetes*, the OR was 1.49 (95% CI 0.86 to 2.59). For *asthma*, OR was 0, as none of the 24 health-equity related projects involved any multi-institutional teams, while for cardiovascular, the OR was 0.88, but like all other topics except *asthma*, was not significant (95% CI 0.45 to 1.74). Finally, compared to R1 institutions, R2 institutions had higher odds of engaging in projects related to health equity. Except for *dementia*, the OR was always higher than 1, but was only significant for *diabetes* (OR=1.89, 95% CI 1.15 to 3.09). Differences between R1 and R2 institutions persisted even when controlling for specific sub-topic e.g., asthma treatment within the asthma topical cluster (Figure 4).

**Figure 4.**
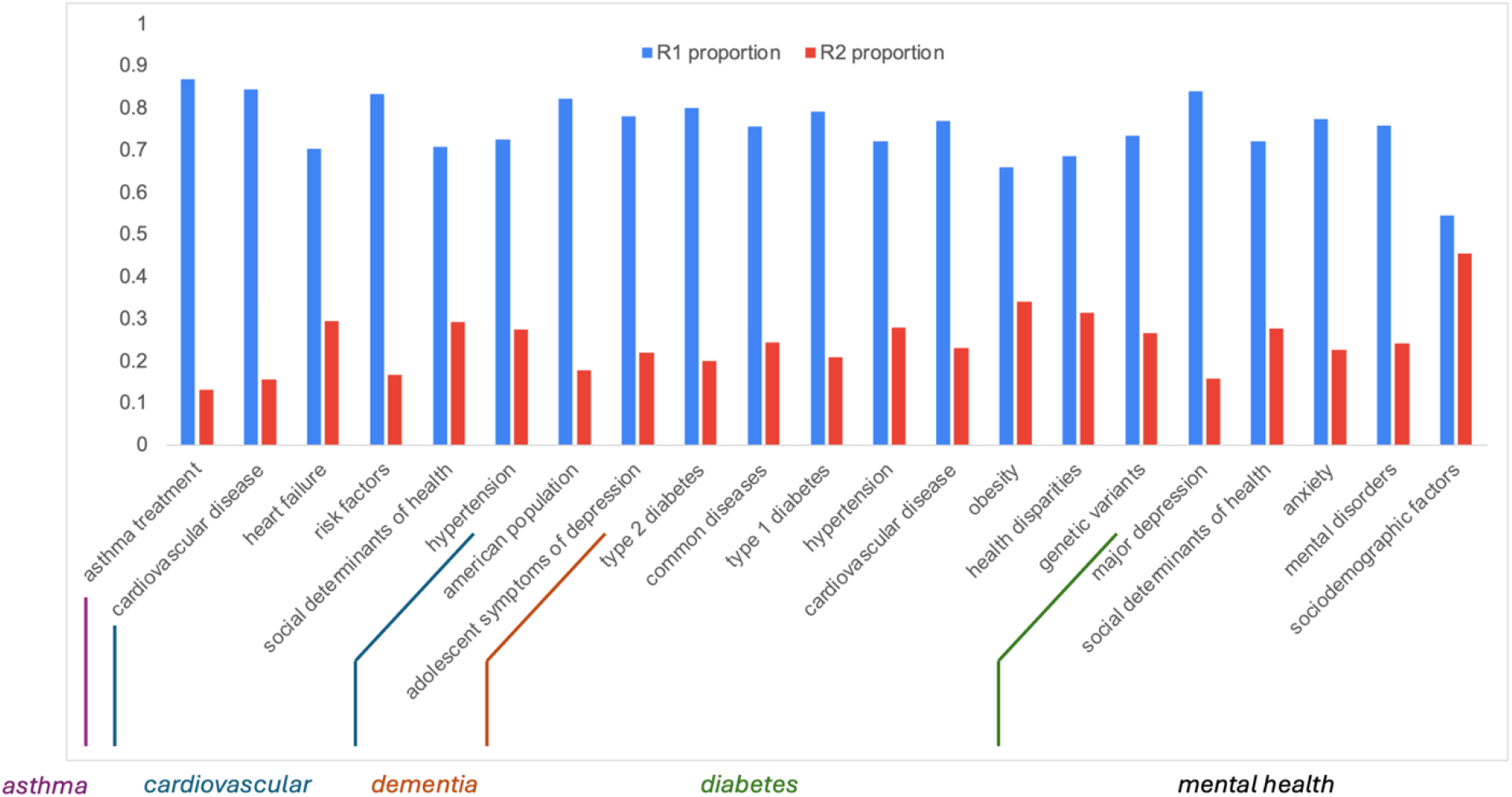
Proportions of R1 and R2-institution projects for the (up to) top 10 highest-frequency keywords across all five topics common to both R1 and R2 projects. With the sole exception of “sociodemographic factors,” which was not significant at the 90% confidence level or above, the R1 proportion was significantly greater than 0.5 at the 99% confidence level or above for all other keywords.

We also performed a qualitative analysis of health equity-related keywords conceptualized from the NIMHD framework. We compared the keywords of R1 and R2 institutions to examine the differences in topic emphasis between these types of institutions. As shown in Figure 5, R1 institutions tend to focus more on topics such as “race/ethnicity,” “socioeconomic status,” “discrimination,” and “sex/gender,” with multiple categories co-occurring these keywords. In contrast, R2 institutions cover a broader spectrum of topics, as evidenced by the greater number of keywords. Specific topics that are unique to R2 institutions include “Hispanic,” “Puerto Ricans,” and “housing instability.” This pattern underscores the differing approaches to health equity research between R1 and R2 institutions, suggesting that the contributions of the latter are important to the broader conversations on promoting and understanding drivers of health equity research. Detailed keyword comparisons for all categories are included as additional data.

**Figure 5.**
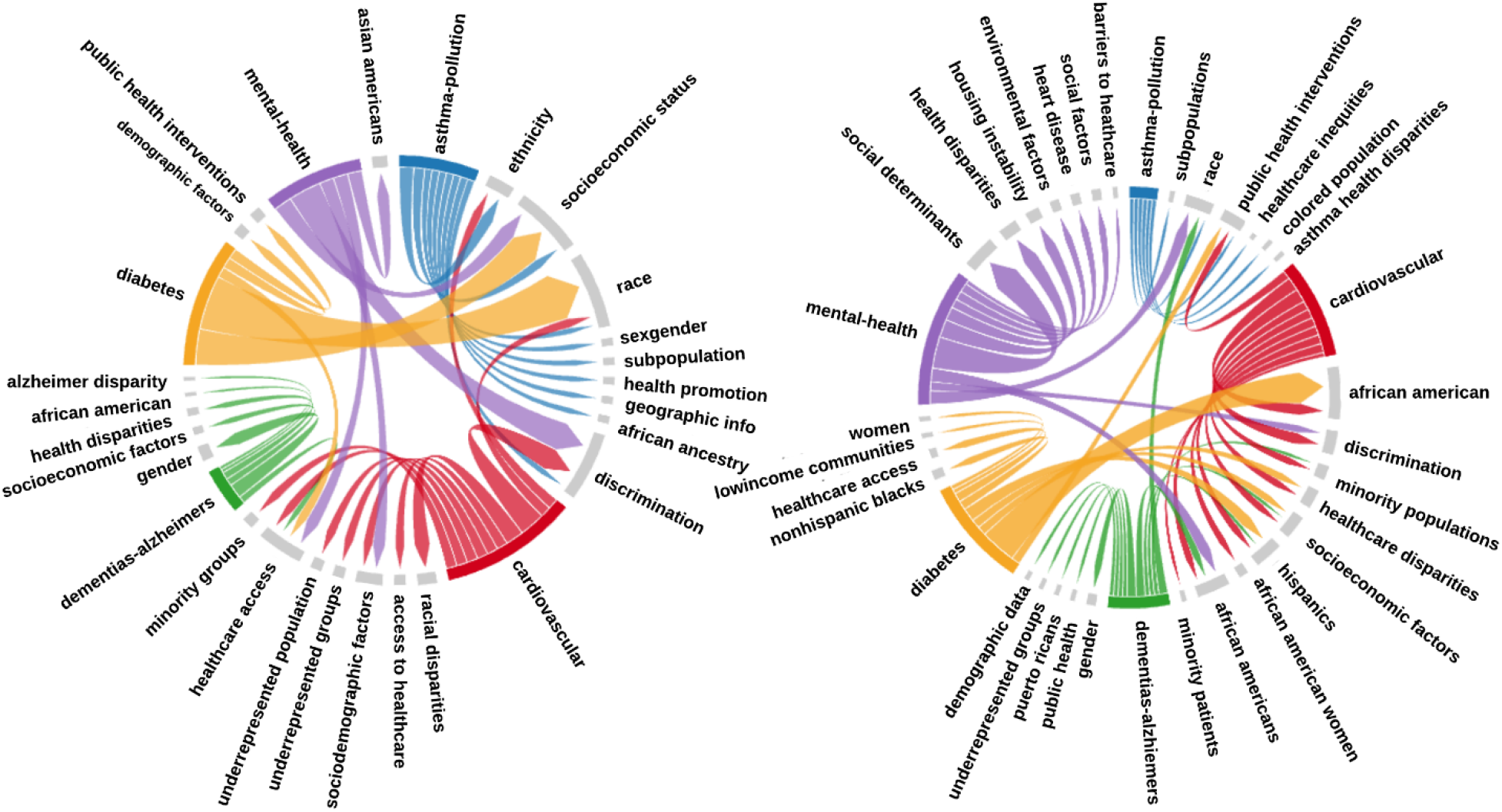
Two chord diagrams illustrating health equity related keywords co-occurrence from R1 institutions (left) and R2 institutions (right). Each band connects two keywords, where the band width is proportional to co-occurrence frequency. Five band colors represent the topics (blue = *asthma*, purple = *mental health*, red = *cardiovascular*, green = *dementia*, yellow = *diabetes*), and the grey band represent *health equity* keywords.

## Discussion

In this study, we demonstrate that GPT-3.5 can perform a range of tasks related to medical information extraction from free text that could enable us to conduct thematic health equity analysis. Starting with the creation of a Python script to extract data from HTML files hosted by the *All of Us Research Hub*, GPT-3.5 was utilized to automate the extraction of institutional affiliations, classify R1 and R2 institutions, and identify health equity-related keywords. These preprocessing steps are fundamental for accurate data analysis in medical research pipelines, reducing manual burden and increasing efficiency of data preparation.

To complement our statistical analysis, we extend the discussion with qualitative analysis using an example that demonstrates the potential of LLM-assisted human analysis in medical research. Specifically, we selected a research project from the category with the highest odds of being associated with multi-institutional collaborations (the mental health category). According to our statistical analysis, mental health projects, as shown in Table 2, exhibited the strongest association with multi-institutional collaborations.

This observation aligns with prior studies supporting collaborations to improve the quality of mental health research. For instance, [28] emphasizes that collaborations among researchers, clinicians, and individuals with mental illness are crucial for producing relevant, feasible, and ethical research. Similarly, [29] explores organizational structures and barriers to collaboration with consumers in mental health research. The authors advocate for a systematic and strategic approach to advance mental health consumer research, highlighting that collaborations are “always worth the extra effort.” Furthermore, several research initiatives show the prominence of multi-institutional collaborations in mental health research [30-32]. This aligns with our statistical findings demonstrating a strong association between mental health projects and collaborative efforts across institutions.

In Figure 6, we present the project titled “Classification of Mental Health Disorders and Social Determinants of Health,” which is related to health equity and conducted by multiple institutions. This example highlights how GPT-3.5 correctly classified Rutgers as an R1 institution and the City University of New York (CUNY) as R2. Additionally, GPT-3.5 processed unstructured fields, such as *Scientific Questions, Project Purpose(s), Scientific Approaches*, and *Anticipated Findings*, converting them into UMLS-coded keywords. The model demonstrated its ability to summarize complex medical terminology from raw text, such as translating “psychiatrist diagnosis” into the UMLS keyword “Mental health disorders.” Furthermore, it was also able to identify health equity-related keywords, with four out of ten—such as “Social determinants of health,” “Adverse experience,” “Neighborhood characteristics,” and “Racial and ethnic identities”—directly related to health equity. This shows that it can significantly reduced the time researchers would have spent on manual thematic analysis and labeling, allowing them to focus on the main statistical analysis.

**Figure 6:**
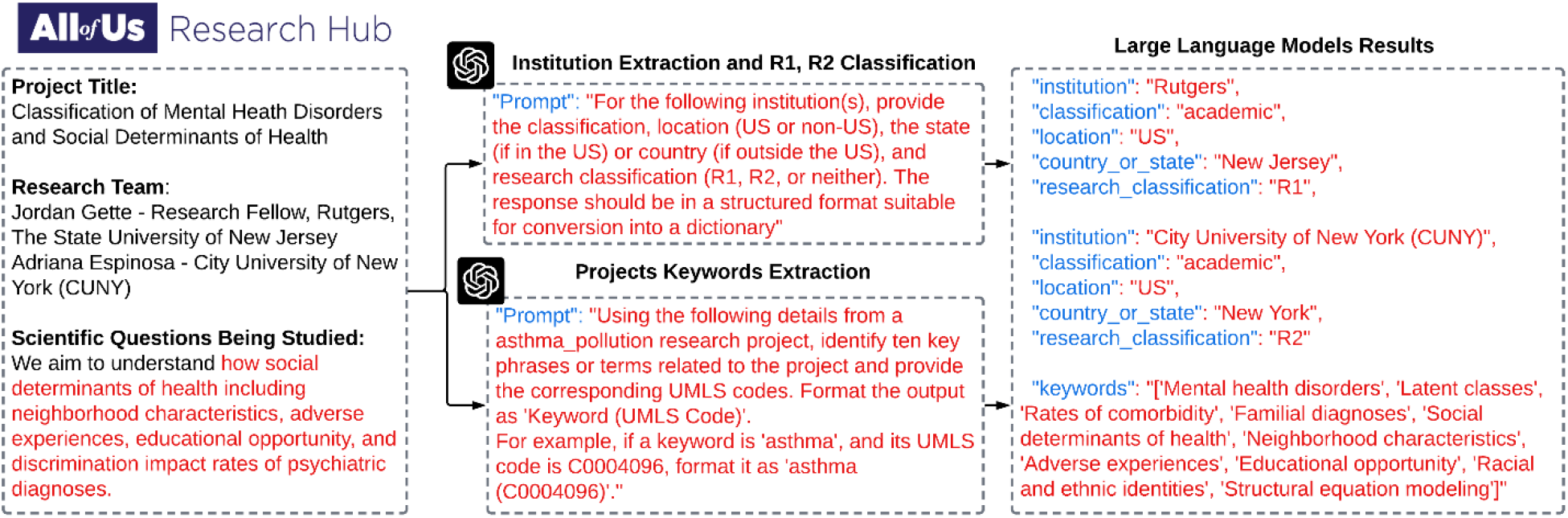
An example of a project from the category with the highest odds of being associated with multi-institutional collaborations (mental health). The figure (left) shows project details related to the analysis, including the research team and scientific questions being studied. Next, the figure (middle) shows how GPT-3.5 prompting can help with information extraction and keyword generation for researchers to get relevant data for the analysis.

### Limitations

Some limitations must be borne in mind when interpreting the study’s external validity. One issue is potential selection bias, since not all institutions and project leaders can make use of *All of Us* (although the program has opened its resources to a broad set of institutions and stakeholders). The dataset is also subject to technical limitations, such as the accuracy of the semi-automatic methods and the GPT-3.5 prompts used. Our preliminary analysis did not provide evidence of *hallucinatory* behavior on the part of the LLM, but some likelihood of it always exists, especially at scale [33-36]. While complete accuracy cannot be presently guaranteed for any automated text analysis, future replication of the analyses here using other LLMs, and topics would strengthen the conclusions.

## Conclusion

A methodological consequence of this study is that it shows that, with judicious use, LLMs like GPT-3.5 and other advanced automated methods make it feasible to study the sociology of projects currently registered on the *All of Us* research platform, especially related to outcomes like health equity. These models enable us to obtain fine-grained data in a relatively unbiased and efficient manner compared to many hours of manual labeling. Our study here used such methods to consider questions at the intersection of health equity, multi-institutional teaming, and the importance of involving and making the *All of Us* platform and data broadly available, and not just to “very high research activity” R1 institutions. Our methods suggest that answering other similar questions at scale may also be feasible, using the recent swath of commercial generative AI models that have become available at relatively low cost. As a policy matter, we also hope that it incentivizes researchers to enter high-quality metadata when registering their projects, as the metadata proves invaluable in conducting such sociological analyses and showcasing the utility of an initiative like *All of Us*.

## Data Availability

All data is publicly available, and linked in the manuscript.

https://github.com/navapatn/all-of-us-projects/blob/main/data/augmented-data/processed_responses.csv

## Acknowledgements

Not applicable.

## Authors’ Contributions

M.K. conceived the study and supervised the work. N.N. conducted the experiments and analysis. Both authors contributed to the writing and revision of the draft. All authors read and approved the final manuscript.

## Abbreviations

AI: Artificial Intelligence
CI: Confidence Interval
CSV: Comma Separated Values
GPT: Generative Pre-trained Transformers
HTML: HyperText Markup Language
LLM: Large Language Model
OR: Odds Ratio
UMLS: Unified Medical Language System
U.S.: United States

## Data Availability

The primary ‘raw’ data that we used is publicly available as *All of Us* project descriptions. The processed and augmented dataset is available here: https://github.com/navapatn/all-of-us-projects/blob/main/data/augmented-data/processed_responses.csv.

## Code Availability

Code for reproducing our analysis (including code for ChatGPT prompt engineering) on the augmented dataset may be found here: https://github.com/navapatn/all-of-us-projects/tree/main/data

For replicating the data augmentation process itself using ChatGPT, we provide prompting examples and codes in these documents: https://github.com/navapatn/all-of-us-projects/tree/main/codes https://github.com/navapatn/all-of-us-projects/tree/main/prompt-examples

## References

1. Braveman P. Health disparities and health equity: concepts and measurement. Annu Rev Public Health 2006;27:167–194.

2. Baciu A, et al. The Need to Promote Health Equity. In: Communities in Action: Pathways to Health Equity. Washington, DC: National Academies Press (US); 2017.

3. Marmot M. Achieving health equity: from root causes to fair outcomes. Lancet 2007;370(9593):1153–1163.

4. Farrer L, et al. Advocacy for health equity: a synthesis review. Milbank Q 2015;93(2):392–437.

5. Braveman P, et al. What is health equity?. Behav Sci Policy 2018;4(1):1–14.

6. Whitehead M. The Concepts and Principles of Equity in Health. Copenhagen: WHO, Regional Office for Europe (EUR/ICP/RPD414 7734r); 1990:29 pp.

7. National Institute on Minority Health and Health Disparities (2017). NIMHD Research Framework. Retrieved from https://nimhd.nih.gov/researchFramework. Accessed on 6/15/2024

8. National Institutes of Health. All of Us: About [Internet]. 2021. Available from: https://allofus.nih.gov/about [accessed 6 Apr 2024].

9. National Institutes of Health. All of Us: Research Projects Directory [Internet]. 2024. Available from: https://allofus.nih.gov/protecting-data-and-privacy/research-projects-allus-data [accessed 6 Apr 2024].

10. Bogard K, Murry VM, Alexander CM. Perspectives on health equity and social determinants of health. National Academy of Medicine; 2017.

11. Embrett MG, Randall GE. Social determinants of health and health equity policy research: exploring the use, misuse, and nonuse of policy analysis theory. Soc Sci Med 2014;108:147–155.

12. Penman-Aguilar A, et al. Measurement of health disparities, health inequities, and social determinants of health to support the advancement of health equity. J Public Health Manag Pract 2016;22:S33–S42.

13. Carnegie Classification of Institutions of Higher Education. Classification Methodology: Basic Classification [Internet]. Available from: https://carnegieclassifications.acenet.edu/carnegie-classification/classification-methodology/basic-classification/ [accessed: 6 Apr 2024].

14. Ostlin P, et al. Priorities for research to take forward the health equity policy agenda. Bull World Health Organ 2005;83(12):948.

15. Rasanathan K, Diaz T. Research on health equity in the SDG era: the urgent need for greater focus on implementation. Int J Equity Health 2016;15:1–3.

16. Thomas SB, et al. Toward a fourth generation of disparities research to achieve health equity. Annu Rev Public Health 2011;32:399–416.

17. Francés F, Parra-Casado D. Participation as a driver of health equity. 2019.

18. OpenAI. Welcome to the OpenAI developer platform [Internet]. 2024. Available from: https://platform.openai.com/docs/overview [accessed: 6 Apr 2024].

19. Lewis SC, Zamith R, Hermida A. Content analysis in an era of big data: A hybrid approach to computational and manual methods. J Broadcast Electron Media 2013;57(1):34–52.

20. Popping R. Analyzing open-ended questions by means of text analysis procedures. Bull Sociol Methodol 2015;128(1):23–39.

21. Van Atteveldt W, Van der Velden MACG, Boukes M. The validity of sentiment analysis: Comparing manual annotation, crowd-coding, dictionary approaches, and machine learning algorithms. Commun Methods Meas 2021;15(2):121–140.

22. Barbosa NM, Chen M. Rehumanized crowdsourcing: A labeling framework addressing bias and ethics in machine learning. In: Proceedings of the 2019 CHI Conference on Human Factors in Computing Systems. 2019.

23. Tecimer KA, et al. Detection and elimination of systematic labeling bias in code reviewer recommendation systems. In: Proceedings of the 25th International Conference on Evaluation and Assessment in Software Engineering. 2021.

24. Richardson L. Beautiful soup documentation [Internet]. 2007. Available from: https://readthedocs.org/projects/beautiful-soup-4/downloads/pdf/latest/ [accessed 6 Apr 2024].

25. Kejriwal M, Miranker DP. An unsupervised instance matcher for schema-free RDF data. J Web Semantics 2015;35:102–123.

26. Bodenreider O. The unified medical language system (UMLS): integrating biomedical terminology. Nucleic Acids Res 2004;32(Suppl_1):D267–D270.

27. Holtz Y. Chord diagram [Internet]. Available from: https://r-graph-gallery.com/chord-diagram.html [accessed 6 Apr 2024].

28. Brandon A. Kohrt, Nawaraj Upadhaya, Nagendra P. Luitel, Sujen M. Maharjan, Bonnie N. Kaiser, Elizabeth K. MacFarlane, Noreen Khan, Authorship in Global Mental Health Research: Recommendations for Collaborative Approaches to Writing and Publishing, Annals of Global Health, Volume 80, Issue 2, 2014, Pages 134–142,

29. Happell, Brenda, et al. “‘It is always worth the extra effort’: Organizational structures and barriers to collaboration with consumers in mental health research: Perspectives of non-consumer researcher allies.” International journal of mental health nursing 29.6 (2020): 1168–1180

30. AlarcónRenato D et al. Hispanic immigrants in the USA: social and mental health perspectives. The Lancet Psychiatry, Volume 3, Issue 9, 860 –870

31. Mongelli, Francesca, et al. Challenges and Opportunities to Meet the Mental Health Needs of Underserved and Disenfranchised Populations in the United States. Focus, Https://psychiatryonline.org/doi/full/10.1176/appi.focus.20190028, vol. 18, no. 1, American Psychiatric Publishing, Jan. 2020, pp. 16–24, doi:10.1176/appi.focus.20190028. Winter 2020.

32. Pearman Ann, Hughes MacKenzie L., Smith Emily L., Neupert Shevaun D. Mental Health Challenges of United States Healthcare Professionals During COVID-19. Frontiers in Psychology. Volume 11, 2020

33. Rawte V, Sheth A, Das A. A survey of hallucination in large foundation models. arXiv preprint 2309.05922. 2023.

34. Xu Z, Jain S, Kankanhalli M. Hallucination is inevitable: An innate limitation of large language models. arXiv preprint 2401.11817. 2024.

35. Ji Z, et al. Towards mitigating LLM hallucination via self reflection. In: The 2023 Conference on Empirical Methods in Natural Language Processing. 2023.

36. Martino A, Iannelli M, Truong C. Knowledge injection to counter large language model (LLM) hallucination. In: European Semantic Web Conference. Cham: Springer Nature Switzerland; 2023.

